# The Ugandan Severe Acute Respiratory Syndrome -Coronavirus 2 (SARS-CoV-2) Model: A Data Driven Approach to Estimate Risk

**DOI:** 10.1101/2020.12.28.20248922

**Authors:** Betty Nannyonga, Henry Kyobe Bosa, Yonas Tegegn Woldermariam, Pontiano Kaleebu, John M Ssenkusu, Tom Lutalo, Willford Kirungi, Fredrick Edward Makumbi, Vincent A Ssembatya, Henry G Mwebesa, Diana Atwine, Jane Ruth Aceng, Rhoda K Wanyenze

## Abstract

**Objectives:** The first case of Severe Acute Respiratory Syndrome-Coronavirus 2 (SARS-CoV-2) was identified on March 21, 2020, in Uganda. The number of cases increased to 8,287 by September 30, 2020. By May throughout June, most of the cases were predominantly imported cases of truck drivers from neighbouring countries. Uganda responded with various restrictions and interventions including lockdown, physical distancing, hand hygiene, and use of face masks in public, to control the growth rate of the outbreak. By end of September 2020, Uganda had transitioned into community transmissions and most of the reported cases were locals contacts and alerts. This study assessed risks associated with SARS-CoV-2 in Uganda, and presents estimates of the reproduction ratio in real time. An optimal control analysis was performed to determine how long the current mitigation measures such as controlling the exposure in communities, rapid detection, confirmation and contact tracing, partial lockdown of the vulnerable groups and control at the porous boarders, could be implemented and at what cost.

**Methods:** The daily confirmed cases of SARS-CoV-2 in Uganda were extracted from publicly available sources. Using the data, relative risks for age, gender, and geographical location were determined. Four approaches were used to forecast SARS-CoV-2 in Uganda namely linear exponential, nonlinear exponential, logistic and a deterministic model. The discrete logistic model and the next generation matrix method were used to estimate the effective reproduction number.

**Results:** Results showed that women were at a higher risk of acquiring SARS-CoV-2 than the men, and the population attributable risk of SARS-CoV-2 to women was 42.22%. Most of the women affected by SARS-CoV-2 were likely contacts of cargo truck drivers at the boarders, where high infection rates were reported. Although most deaths in Uganda were in the age group of 60-69, the highest case fatality rate per 1000 was attributable the age group of 80-89, followed by 70-79. Geographically, Amuru had the highest relative risk compared to the national risk to SARS-CoV-2. For the case of mitigation scenarios, washing hands with 70% com pliance and regular hand washing of 6 times a day, was the most effective and sustainable to reduce SARS-CoV-2 exposure. This was followed by public wearing of face masks if at least 60% of the population complied, and physical distancing by 60% of the population. If schools, bars and churches were opened without compliance, i.e., no distancing, no handwashing and no public wearing of face masks, to mitigation measures, the highest incidence was observed, leading to a big replacement number. If mitigation measures are not followed by the population, then there will be high incidences and prevalence of the virus in the population.

## Introduction

On 30 January 2020, the WHO declared the outbreak of coronavirus disease (SARS-CoV-2) a Public Health Emergency of International Concern. The new virus was highly infectious and transmitted by respiratory droplets when an infected person coughs or sneezes (WHO, Modes of transmission of virus causing COVID-19: implications for IPC precaution recommendations, March 29 2020, Scientific Brief). As of October 19, 2020, 39,801,612 confirmed cases were reported worldwide, including 10,455 for Uganda (WHO Coronavirus disease (COVID-19)/Situation report, 2020), where the first case was reported on March 21. The virus, which emerged out of the city of Wuhan, China, in December 2019 has demonstrated the capacity to generate violent outbreaks even in confined settings and across borders following human mobility patterns (Muzimoto and Chowell, 2020). Although the symptoms are predominantly mild, it can also lead to severe disease especially among the older populations and individuals with underlying health issues (Adler, 2020).

There are four phases as a pandemic evolves in a country. In Phase 1 there are no cases, ports of entry are controlled, and travels to high risk areas restricted. Uganda was in this phase up to March 21, 2020, when the first case was reported. Phase 2 is when sporadic cases arise. The main aim here is to prevent local transmission to contacts, with containment measures including identification and quarantine of suspected individuals/cases. Due to returning residents, sporadic cases were reported in Uganda till around May 09, 2020 when lockdown was eased. In Phase 3, clusters of community spread are observed. After relaxing restrictions, Uganda transitioned from sporadic cases to clusters at ports of entry (PoE) at Elegu/Nimule and Malaba due to cross border build-up by exiting and incoming trucks, and slow testing and delivery of results. The major aim of the national response during this phase was to prevent transmission beyond the infected clusters. Uganda remained in this phase till around May 09, 2020 when the lockdown was eased. Even with the introduction of extensive containment measures, non-adherence to mitigation measures was a major challenge and hence Uganda started transitioning to Phase 4, with widespread community spread. Several indirect local contacts and multiple unconnected clusters of SARS-CoV-2 had been reported by end of September 2020. Deaths continued to rise, and the epidemic curve was oscillating (Figure 1).

**Figure 1:**
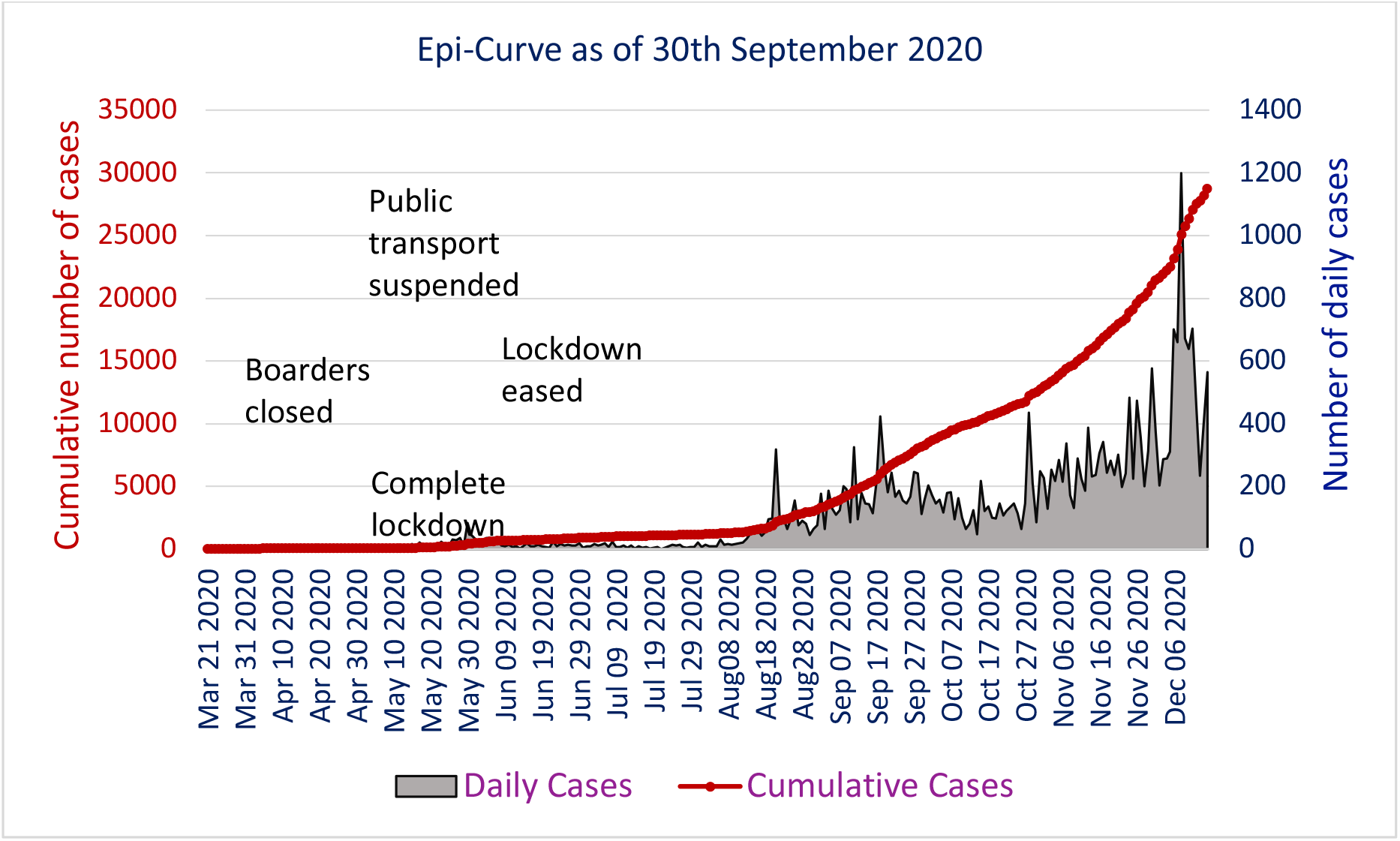
The epidemic curve as of December 15 2020

Since the emergence of clusters, there was re-deployment of efforts targeting the epicentres and PoE, in order to optimize resources. Uganda also enhanced border testing, surveillance and management of cases. In this study we performed risk analysis for Phases 2 and 3, and modelled Phase 4 for to inform planning, in the event of total lifting of lockdown.

The data used in the study was obtained from the daily series of confirmed cases of SARS-CoV-2 in Uganda from March 21 to September 30, 2020 and is readily available from the World Health Organization (WHO, World Health Organization, Uganda, 2020, https://covid19.who.int/region/afro/country/ug), and the Uganda Ministry of Health, MOH – Ministry of Health, Coronavirus (Pandemic) SARS-CoV-2, 2020, www.health.go.ug, @MinofHealthUG). It includes dates of reporting for all confirmed cases, the district of sample collection points, date of confirmation, date of hospitalisation, district of residence, isolation unit and current location (Ministry of Health, Coronavirus (Pandemic) SARS-CoV-2, 2020). For returning citizens, it includes the country of travel and arrival dates in Uganda. For all the confirmed cumulative cases, their close contacts were examined for symptoms and quarantined. The data also indicates whether the cases are local or imported and summarised the clusters comprising one or more cases according to the source of infection. Accordingly, high risk areas were identified for both importation and/or local transmission. The total number of confirmed and exited cases as of September 30, 2020, the case fatality rates, and associated risks as well as the growth rate distribution by gender and age, are presented in Table 1.

**Table 1:**
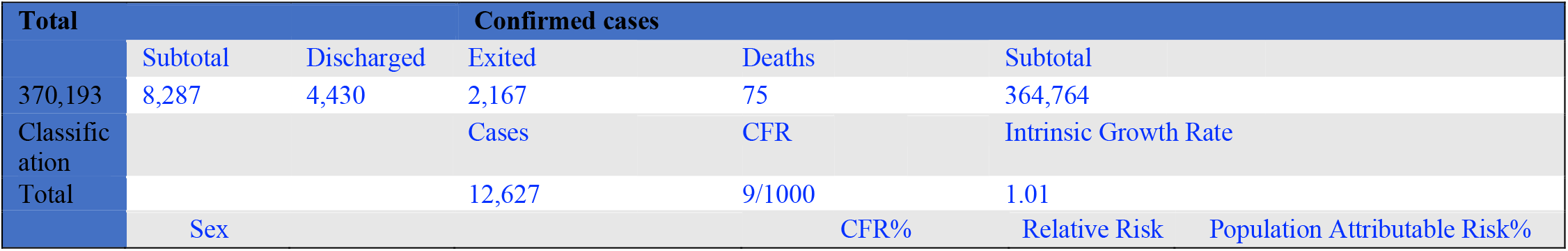
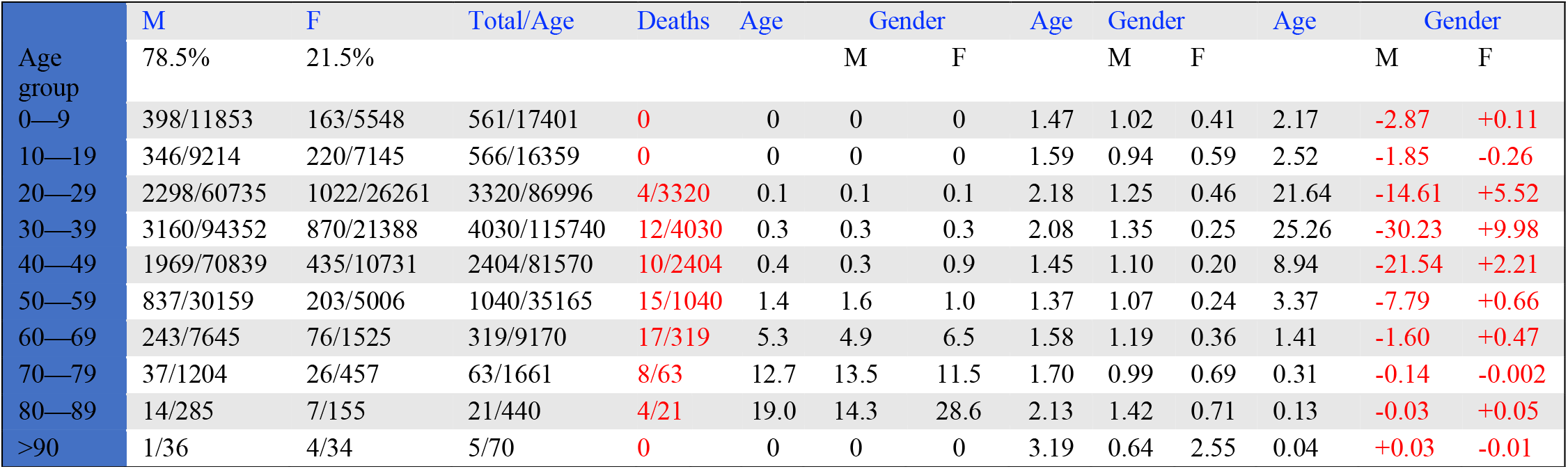
Sample number of confirmed and suspected cases as of September 30, 2020, and distribution by gender and age

## Methods

### The Uganda SARS-CoV-2 model

To model SARS-CoV-2 in Uganda, a flow diagram is used based on characterization of the virus between four states of the susceptible (S), exposed (E), infectious (I), and the removed (SEIR). The susceptible state (S) involves the protected (S_N_, and those with a high risk of exposure S_R_). The infectious class was expanded to include all the confirmed cases of SARS-CoV-2 irrespective of status of infection. These included the quarantined (institutionalized -Q_I_, non-institutionalized, Q_N_), the confirmed mild (M_I_), moderate (M_O_), severe (S_E_) and critical (C). The hospitalized (before recovery) are also collapsed into the infectious state and also incorporates the pre-symptomatic cases irrespective of severity. The removed state includes the dead (D) and recovered. Using the world health organization guidelines (Cabore et al 2020), only a proportion of 3.7% of the entire susceptible population are at risk of exposure. We based our model on these concepts, parameterised using SARS-CoV-2 data from Uganda and determined the parameters that influenced its transmission from March 21 2020, to September 31 2020.

The population was therefore divided into fifteen subgroups referred to as transition states, associated with the pandemic and the current mitigation measures in Uganda. Before the first case was reported on March 21 2020, the country was under partial lockdown, with schools closed but other sectors operational. After registering its first case, a complete lockdown was declared and movement restricted. A call was made to the public for all returnees to report for testing, in addition to their contacts. All those individuals were considered exposed and placed under self or institutional quarantine. While there, confirmation tests were done to determine whether they were infected or not, and if so, the severity of their disease determined. These dynamics and mitigation measures informed the design of the model.

Individuals who were home during the lockdown were label the protected or low risk group (*S*_*N*_). During the lockdown, some individuals left home for essential and emergency duty, and their chances of interacting with suspected cases or exposed individuals was higher than that of the people at home. In addition, health workers continued to test, treat and attend to exposed individuals at the quarantine centres or boarders. We call high risk places epicentres and people who visit such places, the high risk group labelled (*S*_*R*_). All individuals who returned from high risk countries and their contacts were exposed (E), and upon detection and tracing, they either remained under non institutional quarantines (*Q*_*N*_), or were taken to institutional quarantines *Q*_*I*_. Some of these individuals had already been tested but returned home awaiting the test results which later turned out to be positive. Therefore, these individuals were positive while in the community, but did not display any symptoms and had to be removed upon confirmation of positive results (asymptomatic, labeled *A*_*C*_). In some cases, people went to the hospital when they had symptoms (labeled *I)* where all individuals and their contacts were taken upon confirmatory tests, and characterised as either positive but asymptomatic (*A*_*I*_), mild symptoms M_I_, moderate symptoms *M*_*O*_, the severely infected *S*_*E*_, and the critical *C*. The severely and critically infected are the ones that would need hospital bed care and therefore in our model form a basis for threshold number of bed units. All the confirmed cases upon treatment can recover and join class R. Those who do not recover die and join class *D*.

Movement from one group to another implies a reduction (-) from where they are coming from, and an increase (+) to where they are going. Therefore, the system is closed and assumed constant without demographics to keep the population constant. These transitional probabilities are estimated from the MOH data and inputs from the World Health Organization (WHO) (Cabore et. al., 2020). We also assume frequency dependent transmission, where a successful positive contact depends on the probability of infection and the number of contacts made by an individual in a given day. The risk of exposure to Ugandans is 3.7% of the entire population, and only 10% of those result in successful transmission (Cabore et. al., 2020). The 90% are sent to non-institutional quarantine where they return to the population after 14 days. Of the 10% successful exposures, 80% do not develop symptoms while the 20% become symptomatic after an average of 5 days. All the confirmed cases live out their SARS-CoV-2 days in the institutions and only get out after death or recovery. Due to lack of scientific evidence on whether there is re-infection after recovery, in the model we stop at *R* until such information is obtained. The model therefore is an *SEIR* with a segmented *I* class.

With the mitigation measures instituted after the first case in Uganda, the speed of contagion was slowed down, and the policies that greatly lowered the basic reproduction number *R*_*0*_ were containment and suppression. Mitigation measures included quarantine, lockdown, physical distancing, restriction of movement, and public face masking. In view of this, the probability of infection was greatly reduced and this kept community spread to almost 0. Therefore, in order to quantify the mitigation and self-protection measures, transmission rate is assumed to be time-dependent, and a function of the total number of confirmed cases. Although community spread in Uganda was almost zero, the numbers continued to rise due to importation via truck drivers at POE and porous boarders. Thus, we included imports of truck drivers and the undocumented entries at the porous boarders. Once these inflows test positive, their contacts were traced and quarantined. In some cases, the non-citizens were returned to their respective countries. Therefore, the number of cumulative cases in Uganda remained high, but the actual active cases in institutions was less the number returned to their countries. With these assumptions, we presented the transit states for the Ugandan model, and used it to project, predict and come up with different scenarios to inform actions for lifting the lockdown, bed capacity, testing kits, face masks, and treatment procurement.

Movement from one box to another signifies shifts in clinical status of an individual, with the arrow showing where they are leaving and going. As in prior studies on respiratory diseases (Kombe et. a., 2019), we assumed that asymptomatic individuals transmit at a reduced rate compared to those with full blown symptoms. We also assumed that the transmission rate was slowed down by the lockdown, frequent hand washing, public face masks and physical distancing. We therefore constructed a transmission term which is a function of all these mitigation measures. Changing the percentages of these mitigation coverages led to different numbers either applied singly or in combination with others. The best strategy was then identified and forms a basis for actions. In the model, migration was ignored, due to travel restrictions, and also ignored environmental transmission and contamination but instead considered only direct person-to-person infection. These assumptions and descriptions are depicted in Figure 2 and the model given by

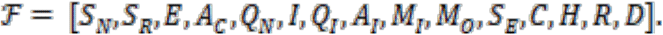

**Figure 2:**
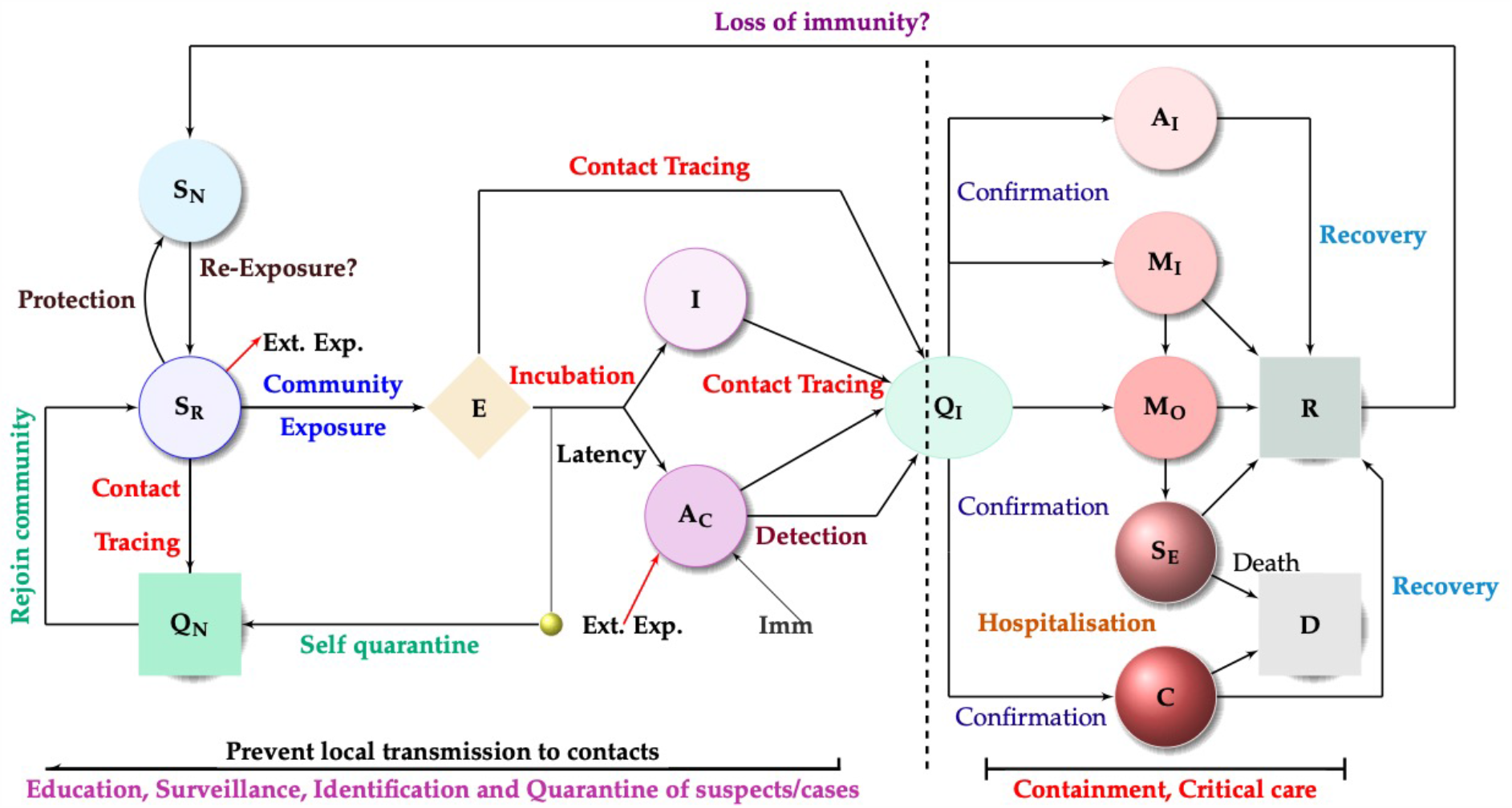
The comprehensive model for SARS-CoV-2

Then, the system of equations to be used is given by

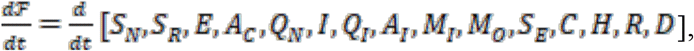

and are the exponential increase for total number of individuals detected and the corresponding contacts traced. The basic reproductive number was determined to be

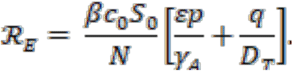

Note that the basic reproduction number is a sum of two terms: the proportion of new infections that arise from the asymptomatic individuals before they shed off the virus, and the proportion from the symptomatic before they are detected.

### The basic reproduction number

The basic reproduction number is the average number of secondary infections that arise from a single infective individual during their infectious period when everyone else in the population is susceptible (van den Driessche and Watmough, 2002; Diekmann et. al., 1990), and depicts the transmission risk in the early phase of SARS-CoV-2 transmission. Due to high transmission rates of the virus, Uganda responded with intensive intervention measures that were expeditiously implemented and led to gradual enhanced self-protection. In order to quantity the daily reproduction number and evaluate the transmission risk changing over time (You et. al., 2020; Nishiura and Linton 2020), the initial contact rate *c*_*0*_ in the expression for the basic reproductive number *R*_*0*_ is replaced by the time-dependent contact rate *c(t)* to reflect the changes of intervention measures and people’s behaviours. This gives

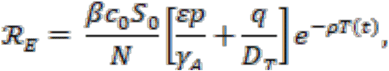

where *ρ* is the level of adherence, and T(t) are the active cases at any time t.

As in prior studies (see Keeling et. al., 2008), in presence of control, the relationship between ℛ_0_ and ℛ_*E*_ is given by

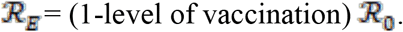

Using this expression we can compute the different levels of adherence with change in the number of active cases in real time.

### Transitional probabilities

Using the WHO guidelines as in Cabore et al 2020, the probability of having mild, moderate, severe or critical disease was approximately 40%, 40%, 15% and 5%, respectively. Although hospitalisation rates depended on policy and capacity, it was estimated that 30% of symptomatic patients needed hospitalisation, with case fatality highest for critical cases (up to 89% without intervention) and 49% for severely ill patients (Oke and Hevenghan). The infection induced mortality rate was estimated at less than 0.1% (Verity et. al., 2109). We also tested our assumptions on parameters using sensitivity analysis, leading to a robust model for infection of SARS-CoV-2 in Uganda.

### Incorporating nonpharmaceutical interventions (NPIs)

The model incorporates the nonpharmaceutical interventions (NPIs) relating to the distancing and hygiene measures Uganda implemented to reduce attack rate. Attack rate is the percentage of an at-risk population that contracts the disease in a specific time interval. The NPIs used were physical/social distancing, and hygiene practices.

For our study, we assumed moderate physical distancing of about 20% and public use of masks of 30% of the population based in the rapid assessment survey. At the time of this analysis, borders and schools were still closed and large gatherings including religious congregations prohibited. There were no interventions to shield vulnerable populations at the time. We however integrated shielded of the vulnerable among the interventions (e.g. shielding the elderly, above 65 yeas, and those with underlying comorbidities such as TB, cardiovascular disease, diabetes among others). We assumed a 20% physical distancing/shielding. We also assumed an increase in hand washing from 26% to 50% as observed from the rapid assessment survey.

### Risk assessment

Risk assessment was conducted and considered a range of factors that might have put different sub groups of people in the population at greater risks from SARS-CoV-2, or have a greater impact of the disease.

Factors considered relevant to assessment of SARS-CoV-2 risk included

- age
- biological sex
- health conditions, and
- location
- nature of exposure (whether import or local alert).

Research has reported that people with specific underlying conditions showed increased risk of severe illness. In addition, being male had also been associated with severe disease. Location of high-risk groups whether at ports of entry of contacts of returning residents was also assessed. These were factored in for a more comprehensive individual risk assessment.

### Forecasting SARS-CoV-2

We forecast SARS-CoV-2 using data and predict the expected number of cumulative cases. We use four approaches namely linear forecasting, the trendline, slope and intercept, the equation of best fit to the data. These forecasts based on statistical and/or mathematical models that aim to predict national and regional numbers of new and total SARS-CoV-2

i. deaths
ii. hospitalizations
iii. cumulative cases

per day for the next 4 months..

The forecasting was done using different types of data, such as SARS-CoV-2 data, demographic data, and mobility data, and employed different methods, and estimates of the impacts of interventions (e.g., hand washing, social distancing, use of face masks). These forecasts were useful as real-time tools to help guide policy and planning. Different approached were used to increase the robustness of forecasts.

## Results

### Data analysis

After the first case in Uganda was detected on March 21, 2020, it was followed by sporadic cases, mainly returning travellers and their contacts. Quarantining of these cases was followed by detection of zero or one case on average in the subsequent weeks as shown in the epidemic curve in Figure 1. As of April 01, 2020 during total lockdown, Uganda had reported 41 cases that increased to 57 by May 04, 2020. However, the number of confirmed cases of SARS-CoV- 2 infection started to increase rapidly from May 09, 2020, following the ease of restrictions. Since then, there has been a steady rise in numbers with a record of 423 cases reported on September 18 (Ministry of Health, Coronavirus (Pandemic) SARS-CoV-2, 2020), leading to steady growth of the pandemic in Uganda. The dynamics of the cases also changed geographically, and in reference to age and gender. By July 12, 2020, the majority of the confirmed SARS-CoV-2 cases in Uganda were male (87 per cent) and 66 per cent (476) of the cases were travellers and cross-border truck drivers from neighbouring Kenya, Tanzania, Rwanda, Burundi, and South Sudan. During this period, Uganda also recorded a rise in local transmissions accounting for 34 per cent (248) of cases, and a shift from sporadic cases to clusters of community spread. Hotspots included the border districts with high-volume PoE from South Sudan, Tanzania, and Kenya, as well as Kampala. More than 1,500 people, including 62 children, were under institutional quarantine in 68 facilities across 52 districts. The Ministry of Health (MoH) reported 494 confirmed cases of SARS-CoV-2 including 17 children admitted to 15 designated isolation and treatment facilities. The majority of the cases were asymptomatic or had mild or moderate disease. By August 31, 2020, 76.2% of the cases were male, signifying an increase in female cases. Further, local cases increased from 34% to 71.6% and imported cases reduced from 66% to 28.4%. As of September 30 2020, 78.5% of confirmed cases were male, and 21.5% female. The highest number of cases were in the age group of 30-39 (4,030), followed by 20-29 (3,320) and 40-49 (2,404). This implies that of the total cases of 12,627 confirmed cases, the three age groups accounted for more than 77% of the confirmed cases. However, the highest percentage of deaths were observed in the age group of 80-89 followed by 70-79. This confirmed prior studies (Adler, 2020), that although the highest number of cases were observed in the younger population, the highest number of deaths were in the older population.

The total number of SARS-CoV-2 cases in Uganda as of August 31, 2020 was 3,037 with 1,652 active cases 1,469 recoveries, and 32 deaths. A total of 381,479 samples had been tested, and 2,115 truck drivers exited to their countries (Ministry of Health, Coronavirus (Pandemic) SARS-CoV-2, 2020). Within a month from August to September, the cumulative local cases had risen to 8,287, with 3,782 active and 75 deaths. This signified an increase of 173% for cumulative cases, 129% increase for active cases, and 134% increase for COVID-19 induced deaths.

By the end of August 31, 2020, most of the cases were reported from Kampala (1,008), Amuru (593), Tororo (248), Kyotera (133), Wakiso (123), Namisindwa (89), Buikwe (67), and Luweero (64). Other districts with more than 20 cases included Arua 56, Kikuube 54, Gulu 50, Busia 42, Kitgum 33 and Adjuman 32. As of September 30 2020, many districts had registered very high number of cases compared to previous reports. For example, Kampala had 3,835, with a 280% increase. However, the districts that registered more than 100 cases, also had high percent increase when we compared cases in August and September. These included in order from the highest to lowest, Kapchorwa, followed by Pader, Moroto, Jinja, Mbale, Busia, Gulu, Lira, Masaka, Kitgum, Mukono, Wakiso, Arua, Kampala, Kiryandongo, Tororo, Adjumani, to name but a few. Among all districts, only Amuru and Elegu registered 19% and 53% reductions overall, signifying a 1% and 2% respective case reduction per day.

The gradual lifting of the national lockdown was accompanied by intensified communication of the Presidential directives on the mandatory use of masks in public places and the observance of physical distancing measures. The Government of Uganda also launched mass mask distribution across the country, targeting all individuals aged 6 years and older. By July 21, 2020, 7,121,246 masks had been distributed. Hot-spot districts were prioritized and supported with intensified surveillance, contact-tracing, and testing alongside enhanced community engagement. As of August 31, 2020, the presidential directives had reduced, and this was reflected in the reduced compliance to mitigation measures and noncompliance to mandatory public wearing of facemasks. As of September 30, 2020, wearing masks is sometimes enforced, but in other areas appearing with a mask is optional and observed by non-locals.

Using the nonpharmaceutical interventions relating to the distancing, facemasks, and hygiene, the percentage coverage of each used singly or in combination with others is determined.

### Case fatality risk

Case fatality rate or risk is the proportion of people who have died from SARS-CoV-2 among all individuals diagnosed with the disease over a certain period of time. It is used as a measure of disease severity and for prognosis (predicting disease course or outcome), where comparatively high rates are indicative of relatively poor outcomes. For the case of Uganda, in the sample of 3,037 with SARS-CoV-2, 32 fatalities were reported on August 31, 2020. Thus, the overall case fatality was about 11 per 1000. As of September, the case fatality rate was 9 per 1000 signifying a slight reduction in death rates.

### Age and gender specific risk

Considering gender and age risk to SARS-CoV-2, as of August 31 2020, we found that in age groups of 0-9, 30-39, 40-49, 50-59, 60-69, 70-79 and >90, the risk among women was increased by exposure. Most of the women affected by SARS-CoV-2 were likely contacts of cargo truck drivers at the boarders, where high infection rates were reported. Therefore women in these age groups were more likely to acquire SARS-CoV-2 than the men. On the other hand, men in the age group 10-19, and 20-29 were more at risk of exposure to SARS-CoV-2. Population attributable risk (PAR) is the proportion of the incidence of SARS-CoV-2 in the population (exposed and unexposed) that is due to exposure. It is the incidence of SARS-CoV- 2 in the population that would be eliminated if exposure were eliminated. Calculating the population attributable risk percent allows us to determine what percent of infections due to SARS-CoV-2 could possibly be prevented if high risk gender groups were removed from the population. Comparing these results with the data of September 30, 2020, we found that men were exposed more than the women, the case fatality rate was higher for women that the men. This verifies a gender-based risk for SARS-CoV-2, and gender-based policies are paramount to protect this vulnerable group. For example, in the age group of 80-89, the case fatality rate for females is twice that for the males.

### Geographical risk

In August 2020, Amuru had the highest geographical risk, followed by Luweero, Kampala, Kyotera 8%, Tororo, and Namisindwa. As of September 31 2020, Elegu had an exorbitantly high risk, about q0 times more than the second district of Moroto, Amuru, Tororo, Kampala, Busia, and others as seen in Table 2.

**Table 2:** Intervention scenarios analyzed in the Ugandan model

| Scenario | Description |
| --- | --- |
| 1. Unmitigated | No governmental intervention applied |
| 2. 20% physical distancing | Assuming moderate physical distancing measures are in place with moderate impact (20%) restricting movement or limiting contacts with others |
| 3. Shielding of the Vulnerable | The elderly (>65 years) and those with underlying conditions such as, TB, Diabetes |
| 4. 30% public wearing of face masks | We assumed 30% compliance based on the 2020 rapid assessment survey (RAS) undertaken |
| 5. 50% hand-washing, 6 times a day | We assumed increase in hand-washing from 26% (Uganda Demographic Health Survey 2016) to 50% based on the 2020 online survey by Makerere School of Public Health |

**Table 2:** Mitigation coverages needed to control COVID-19

| Mitigation (Detection rate 5 days),<br>number of contacts = 30 | % coverage | No. of secondary<br>infections<br>$R_0$ | Coverage to reduce<br>$R_0$ to less than 1<br>(Thresholds) |
| --- | --- | --- | --- |
| **Used as standalone** |  |  |  |
| Face masks | 30% | 1.46 | 60% |
| Washing hands 6 times a day/sanitizing | 26% | 2.02 | 70% |
| Social distancing | 10% | 1.88 | 60% |
| **Used in combination** |  |  |  |
| Face masks & Washing hands 6 times a<br>day/sanitizing | 30%, 26% | 1.46 | 50%, 30% |
| Social distancing & Washing hands 6<br>times a day/sanitizing | 10%, 26% | 1.88 | 50%, 40% |
| Face masks & Social distancing | 30%, 10% | 1.32 | 60%, 50% |
| **Used simultaneously** |  |  |  |
| Face masks & Washing hands 6 times a<br>day/sanitizing & Social distancing | 30%, 26%,<br>10% | 1.32 | 50%, 30%, 50% |

**Table 3:**
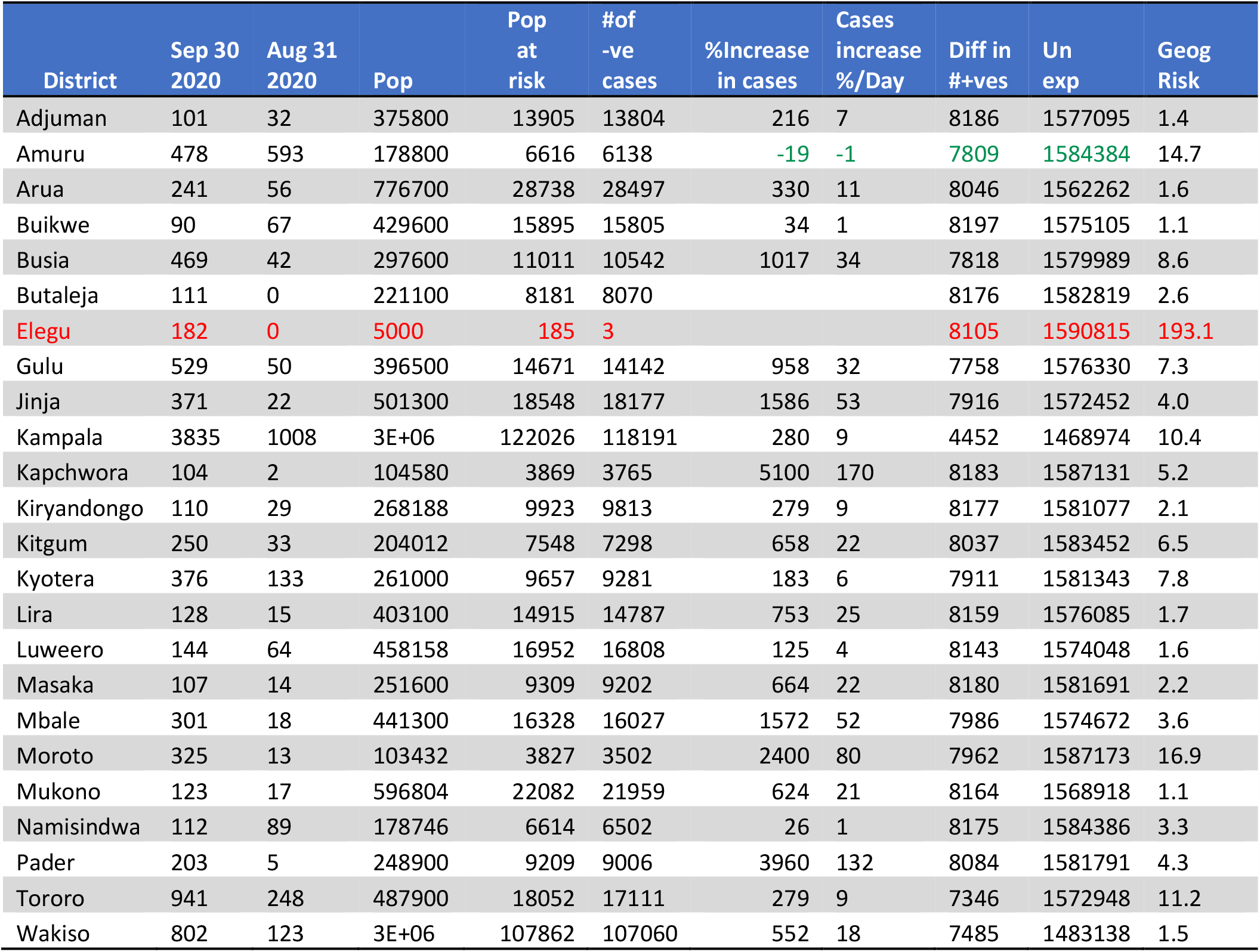
Geographical risks associated with Ugandan districts with more than 100 confirmed cases as of September 30 2020

Although Amuru, Kyotera, Tororo and Buikwe districts had been considered hotspots for SARS-CoV-2 in July 2020, by September 2020, other districts had more confirmed cases although reported lower risk due to the density per square mile which is higher in some districts than in others. Therefore, Elegu for example, the ratio of positive to negative is 1:1, while that in Kampala was 1:1,637 in August, and 1:782 in September. It is important to note however that this applies to Kampala metropolitan, as different and more severe results may be experienced in Kampala city due to a high density of 24,276 per square mile.

### Risk due to imported cases

We observed that although the difference between the local and imported cases was almost twice the local cases, the risk due to imports is gradually increasing from 0.08 in July, 0.13 in August, and 0.27 in September 2020. Therefore, with more imports, the attack rate increases, and mitigation measures to detect imports at points of entry should be put in place.

**Table 4:** Estimating attack rate due to local/imported cases

| Estimating attack rate due to imported cases | July 12, 2020 | August 31, 2020 | September 30, 2020 |
| --- | --- | --- | --- |
| Local cases only | 1,006 | 3,037 | 8,287 |
| Total cases (imported + local) | 2,246 | 5,039 | 12,627 |
| Reported imported cases | 1,240 | 2,002 | 4,340 |
| Ratio of Local: Imported | 1: 1.2 | 1.5: 1 | 1.9: 1 |
| Local attack rate % | 0.06 | 0.19 | 0.52 |
| Total attack rate (imported + local) % | 0.14 | 0.32 | 0.79 |
| Increase in attack rate $\Delta\%$ | 0.08 | 0.13 | 0.27 |

The question to be answered was, since Uganda had transitioned to local clusters, what was the best strategy to mitigate the rising number of cases. We used a linear exponential model to compare to data, repeated the procedure for a nonlinear exponential and later a logistic model. We later on used the data to fit to a comprehensive deterministic model incorporating all the mitigation measures undertaken by the government of Uganda. In July, the ratio of local to imported was 45%, in August it had increased to 60%, and continued to increase to 66% by September 2020. This implies that more local transmissions are reported and continue to increase compared to July when most if the cases were imported. Therefore, Uganda transitioned to Phase 4 of COVID-19 transmission, with widespread community spread.

### Projecting using linear exponential, nonlinear exponential and the logistic model

We forecast SARS-CoV-2 using data and predict the expected number of cumulative cases as shown in Figure 3. Figure 3 gives the number of actual cumulative cases, forecasted cumulative cases and the undetected. The figure shows that the actual reported and forecasted cases are almost equal with a few exceptions when the model over or underestimates the cases. When the forecasted numbers are greater than the actual, this would imply un detection of positive cases, while a forecast less than actual would signify a situation when positive contacts of confirmed cases self-isolate till the virus clears.

**Figure 3:**
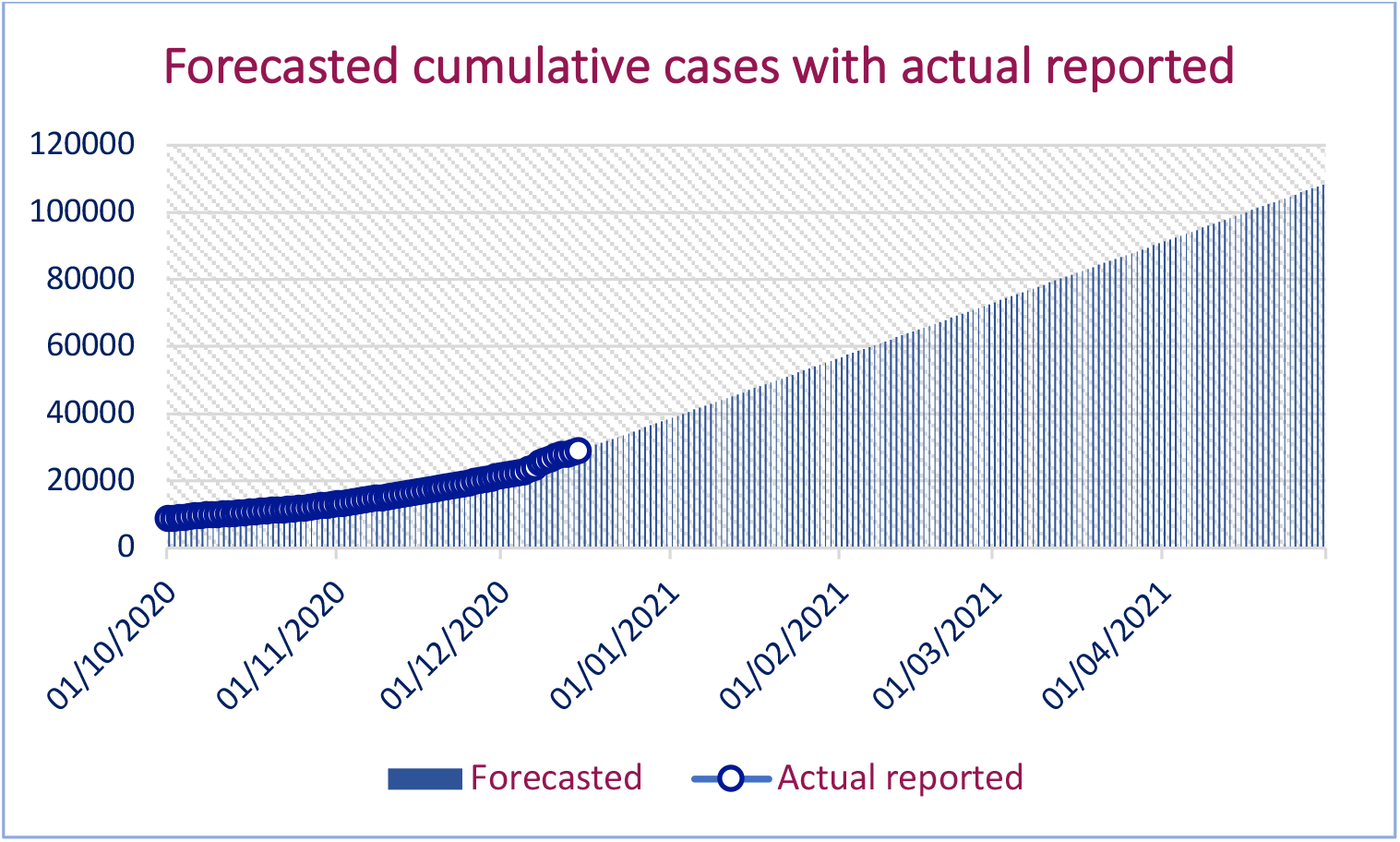
Forecasting COVID-19 up to April 30 2021

The number of undetected and in self-isolation is shown in figure 4.

**Figure 4:**
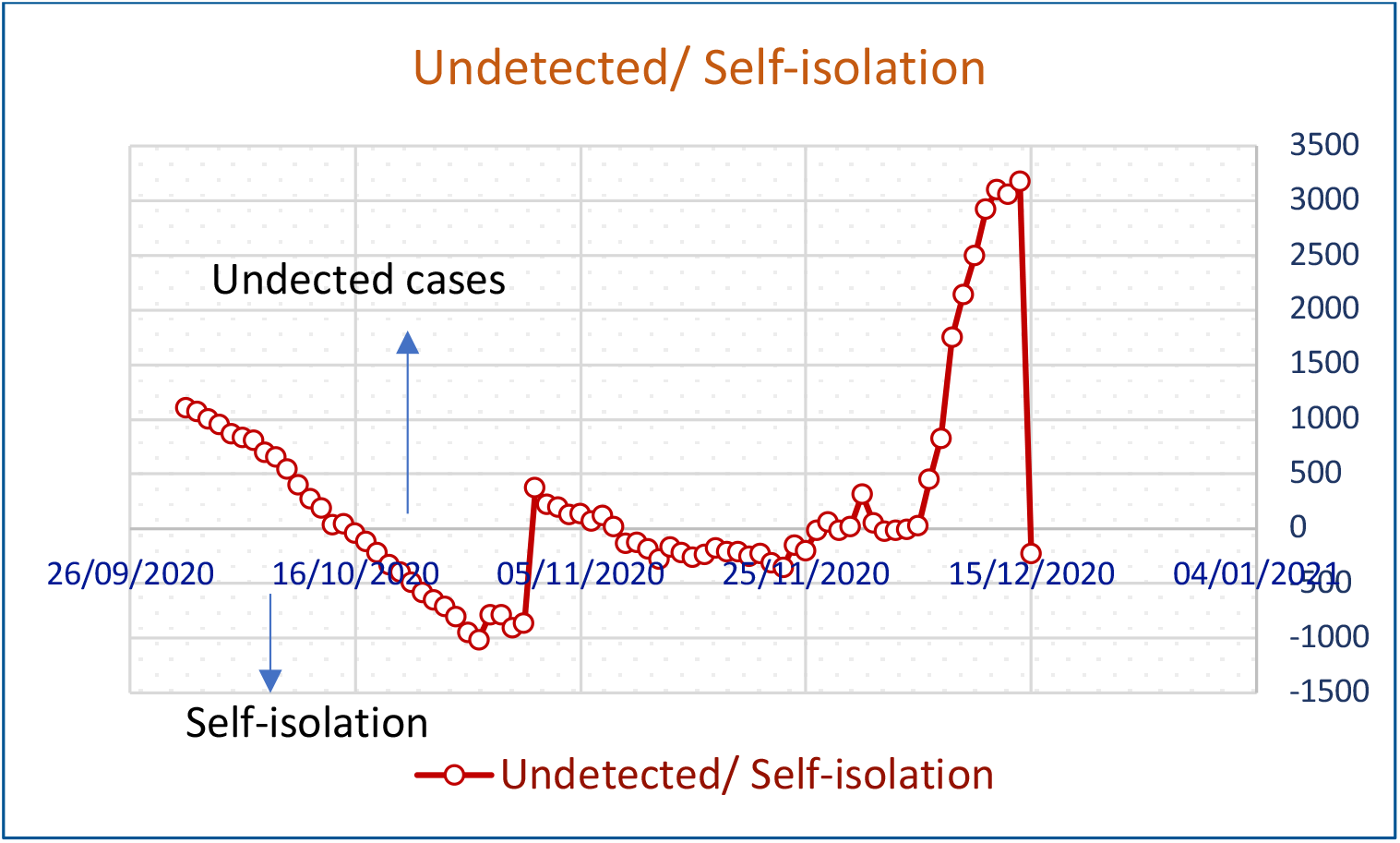
Undetected and self-isolation COVID-19 cases up to April 30 2021

We see that there are more undetected cases than those in self-isolation. This figure would also imply that the cases that could be in isolation are actually not self-isolating but freely mixing with the unsuspecting population.

### Forecasted deaths

Using the deaths cases reported, we forecast the expected number of deaths, updated monthly from the first deaths on July 25, 2020. It is observed from the data that more than 41% of the deaths occurred in the month of November and so far about 9% deaths in December. Using the November deaths, we obtained the upper bound of the projected deaths while December gave the lower bound as shown in Figure 5. All the months fell within this interval.

**Figure 5:**
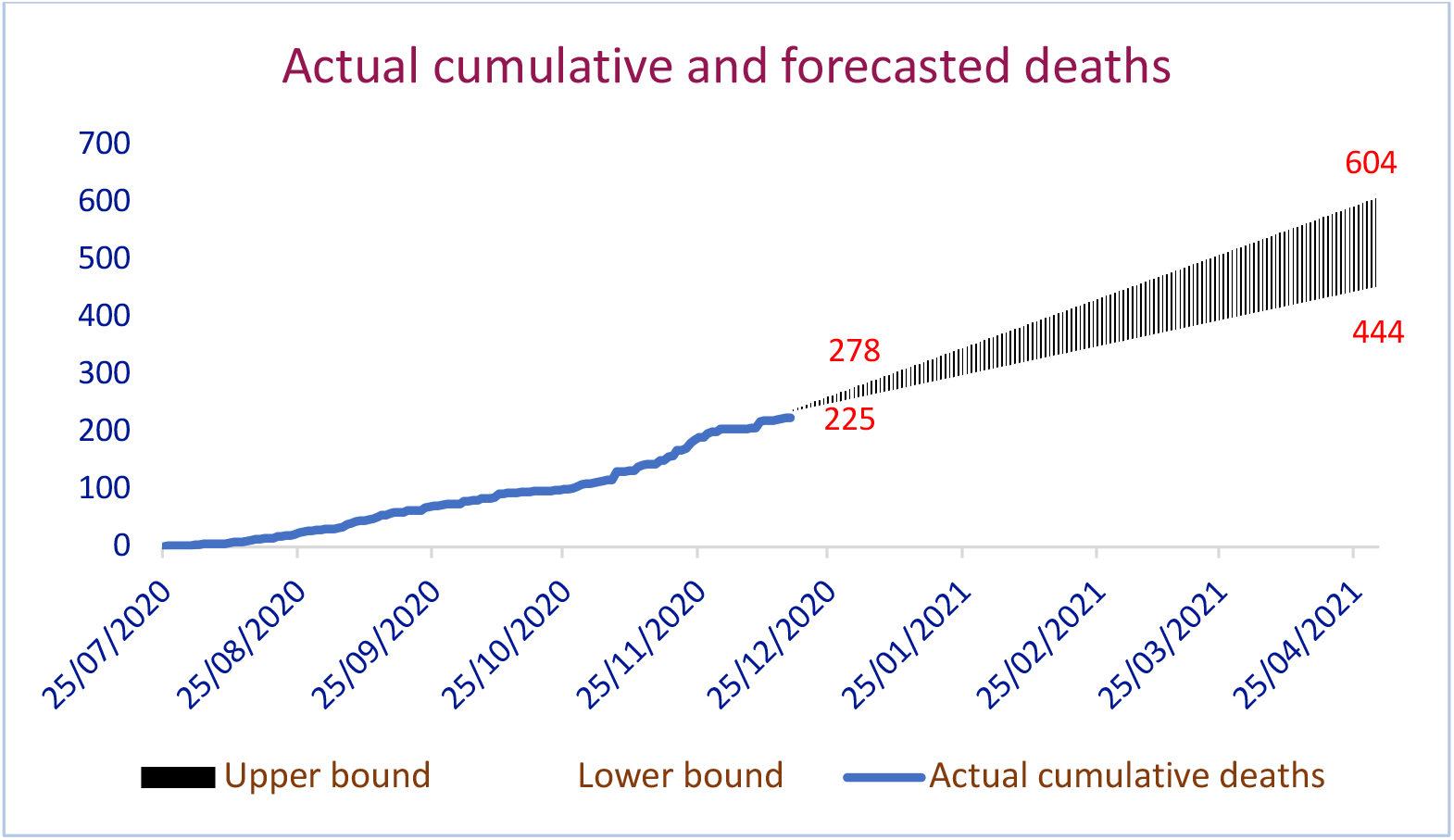
Forecasted deaths

### Vaccine coverage needed to control the virus

The data was also used to calculate the vaccine coverage needed to attain herd immunity. Using different disease severity of asymptomatic, mild, moderate, severe and critical, the basic reproductive number was calculated and from it, a corresponding vaccine coverage obtained with different vaccine efficacies.

It is observed from Figure 6 that high vaccine coverages are needed whenever most of the confirmed cases are critical, while low vaccination levels are obtained for asymptomatic cases.

**Figure 6:**
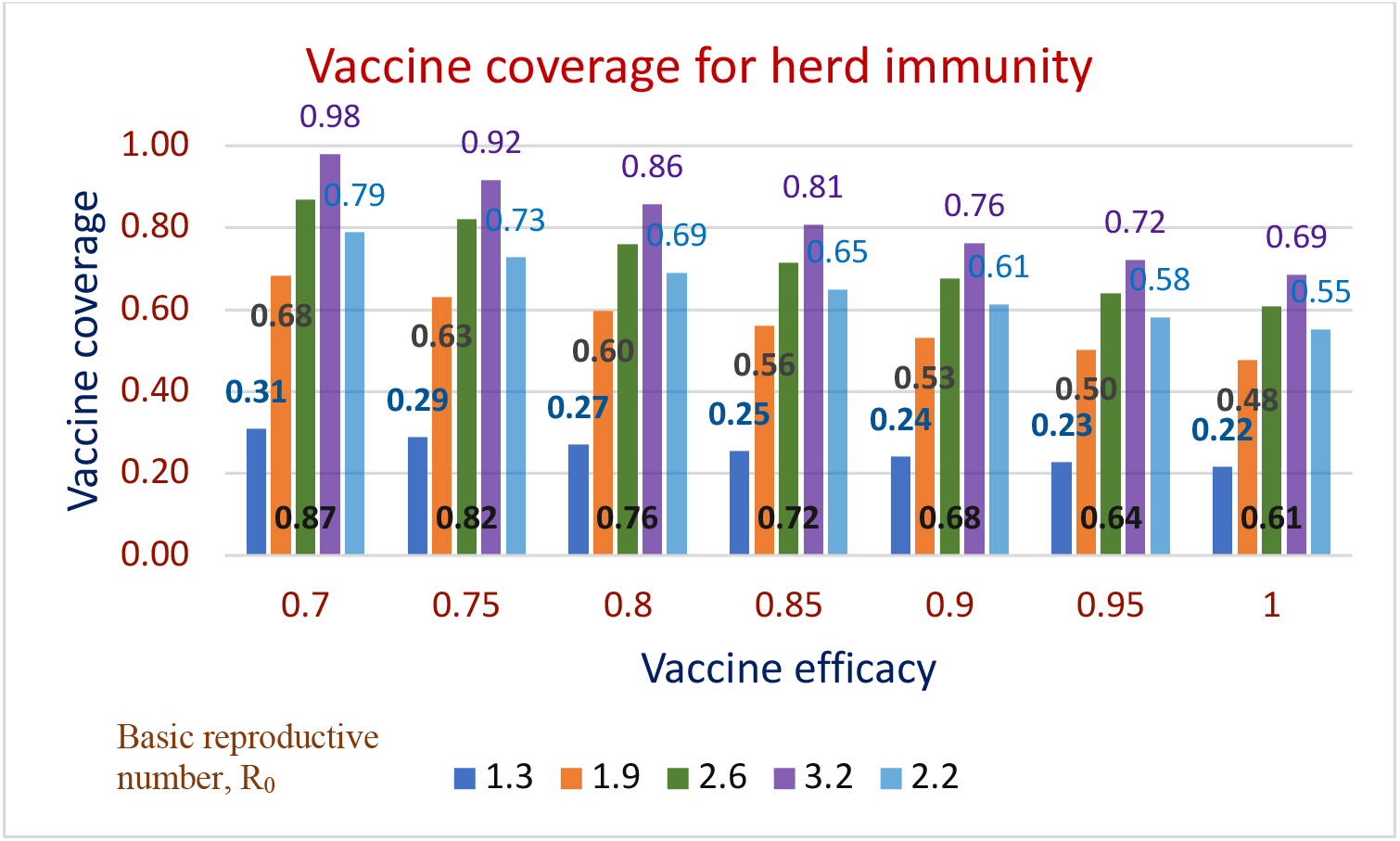
Vaccine coverage given by R_0_

### Sensitivity analysis

Performing sensitivity analysis on the basic reproduction number to determine how the parameters affect ℛ_0_ gives Sensitivity indices help to determine which parameters are responsible for persistence of disease in the population. We observe from the indices that the probability of infection is most sensitive and therefore measures to prevent infection should be enforced. This includes the non-pharmaceutical interventions, and vaccination to reduce susceptibility of the population. Reducing susceptibility implies targeting those populations that are vulnerable to prevent exposure. The recovery rate is also very sensitive to how many secondary infections are expected from a single exposure. The longer the recovery time is, the more likely a secondary infection. Preventing such infections would include vaccination of health personnel, and also measures that ensure quick recovery of infectious individuals. The proportion of asymptomatic individuals and their probability of transmission also leads to increase in secondary infections. The higher these values are, the more infections expected. It is therefore imperative that asymptomatic individuals prevent further transmission by observing the set standard operating procedures. The number of symptomatic individuals in the population increases chances of new infections but not as in the asymptomatic case. This is because these people have been identified and can be avoided unlike in the previous cases when an individual might not tell the status of another due to absence of symptoms. The detection rate affects the number of new infections in that when it is high, R_0_ is low, and when it is low, R_0_ is high. Therefore, the longer it takes to detect and confirm a positive case implies that this person can infect more people thereby increasing cases within communities.

**Table 5:**
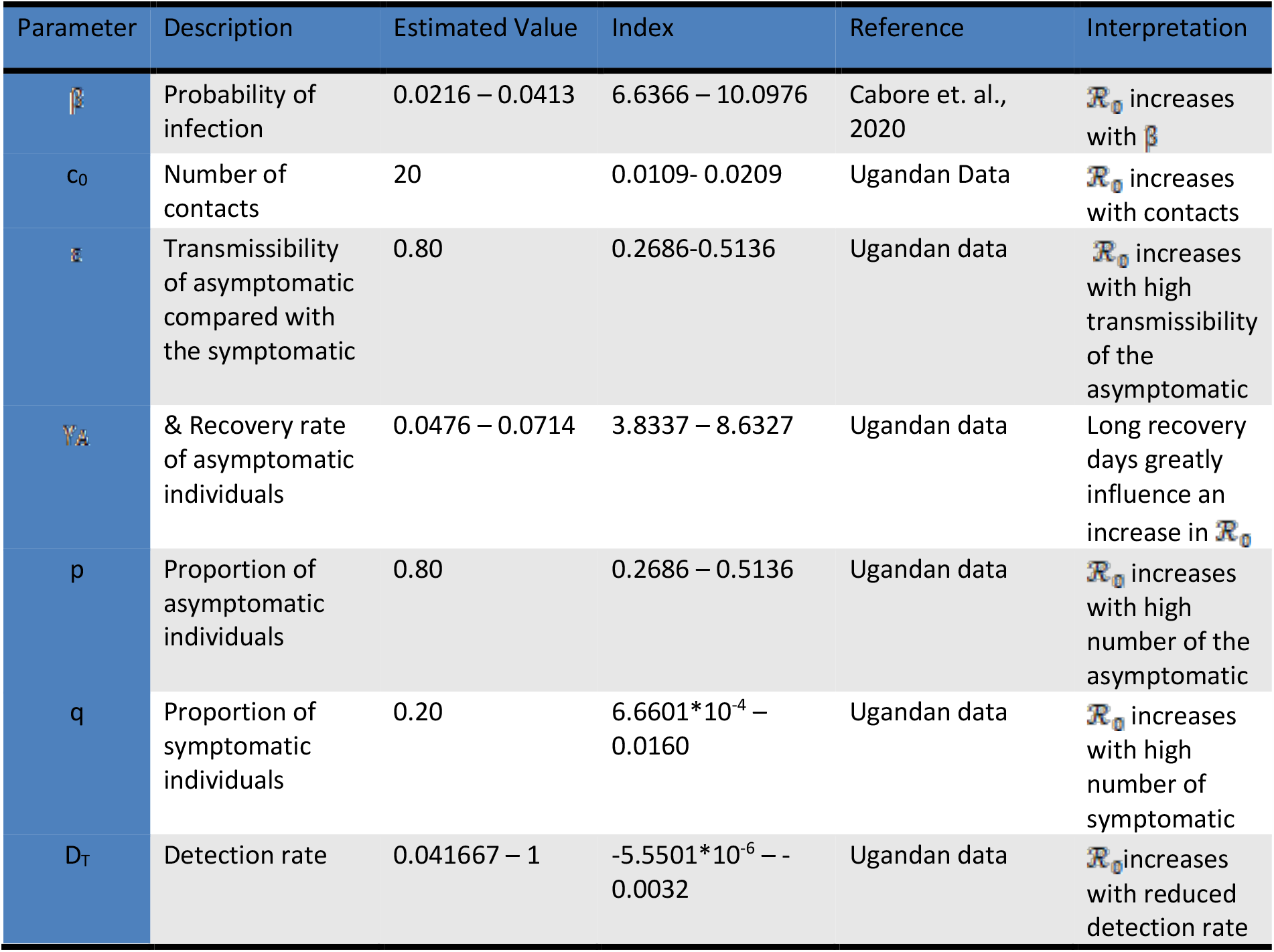
Sensitivity indices of ℛ_0_ parameters

In the preceeding sections, we looked at specific intervention for each of the parameters that influence SARS-CoV-2 in Uganda. The scenarios considered in the model and their descriptions were summarised in Table 2. The full model is simulated with these scenarios and the total cases forecasted within 1 and 2 years are given, with the percentage change in relation to the unmitigated. We observe that apart from shielding the vulnerable groups, the percentage reduction in forecasted cases is more than 30% signifying the constraints of shielding the vulnerable. The most effective scenario is regular washing of hands, followed by wearing masks, and social distancing. We also find that the most consistent scenario to mitigate SARS-CoV-2 is hand washing. For all the scenarios, we show the forecasted number of hospitalised cases. The figure shows that hand washing and wearing face masks in public are very effective to keep the cases low and wipe out the virus. The forecasted cases from the epidemic model are shown in Table 6.

Amidst the growing burden, we modeled the epidemic to inform national planning efforts using four approaches including linear exponential, nonlinear exponential, logistic and deterministic models. We examine different scenarios, where 3.7% of the population are at risk of exposure (Cabore, Karamagi Kipruto et. al., 2020), when the elderly are shielded, schools and churches are open or not, with various compliance levels of physical distancing, hand washing, and examine possibility of allowing social gatherings. The modelling was not intended to reproduce right or wrong answers, and the numbers produced by this model were not presented as exact predictions. However, they provide a planning framework for policymakers by considering various scenarios and intervention strategies and their impact on the epidemic. The data/inputs and assumptions were based on the current number of suspected and confirmed cases of SARS-CoV-2 from March 21, 2020 to August 31, 2020.

## Discussion

Since the first case of SARS-CoV-2 was reported in Uganda on March 21, 2020, the number of cases increased to 1,154 by July 30, 2020. In this study, the risks associated with SARS-CoV-2 in Uganda were estimated and estimates of the reproduction ratio in real time presented.

We identified the high-risk areas and populations, and also considered the drivers of the epidemic, to quantify burden of the disease attributable to a particular risk. For example, the data used showed that the growth of the virus in the country increased whenever high number of truck drivers were reported. To respond to the mounting number of cases, the Ugandan government recommended universal public wearing of face masks, and free issuance to all citizens above 6 years of age. Uganda has also implemented comprehensive physical distancing measures including enhanced infection control measures in hospitals, restricting public transportation, suspension of social events, church services, and delaying the start of school activities (MOH, Ministry of Health: Coronavirus (Pandemic) SARS-CoV-2, 2020). We noted however that due to population densities in urban areas and large house holds, physical distancing was almost impossible to achieve, and could only be achievable with total lockdown, which was not a realistic option for Uganda. With this information, we determined and quantified the risks associated with noncompliance and non-adherence of imposed mitigation measures.

Our results indicate an early sustained transmission of SARS-CoV-2 in Uganda and supports implementation of preventive measures such as physical distancing, wearing face masks in public, and frequent hand washing to rapidly control the outbreak. Results showed that women were at a higher risk of acquiring SARS-CoV-2 than the men, with a population attributable risk of 42.2%. Most of the women affected by SARS-CoV-2 were likely contacts of cargo truck drivers at the boarders, where high infection rates were reported. This scenario was felt in East Africa where fear of COVID-19 slowed trucking in the region (Voice of America COVID-19 Pandemic. Fear of COVID-19 Slows Trucking in East Africa, May 21 2020; Quartz Africa. East African truck drivers carrying essential goods cross-border may also be transmitting Covid-19; The Independent. COVID-19 News. Cargo trucks stuck at borders as countries reject COVID-19 results from EAC bloc). This picture could change over time as Uganda transitions from imported cases to widespread community transmission. In that case, community transmissions may outnumber the imported cases from truck drivers. Our results also supported rapid detection and confirmation as a great reduction to exposure. There is potential for the disease to spread into the community, underscoring the need to implement a wide array of physical distancing measures to rapidly contain the outbreak in Uganda, mitigate the morbidity and mortality impact of the disease, and stem the number of case exportations to neighbouring countries.

Within communities, it is also known that the highest incidence of SARS-CoV-2 is the young population, however, high deaths rates were in the vulnerable group of above 65 years. Severe illness from SARS-CoV-2 increases with age, with older adults at highest risk (Muzimoto and Chowell, 2020; Muzimoto et. al., 2020; Centers for Disease Control CDC, Older Adults. Coronavirus Disease 2019 (COVID-19)). Severe illness means that the person with SARS-CoV-2 may require hospitalisation, intensive care, or a ventilator to help them breathe, or they may even die. For the case of Uganda, the highest number of cases were in the age range of 0- 9, followed by 10-19, and 30-39, (see Table 1. Data extracted from (Ministry of Health, Coronavirus (Pandemic) SARS-CoV-2, 2020).

Although the deaths in Uganda were in the age group of 30-39 and 80-89, the highest risk attributable to age was in the age group of >90, followed by age group 10-19 and by 0-9. Geographically, Kyotera had the highest relative risk compared to the national risk to SARS-CoV-2. In absence of compliance, and with such a high risk of exposure, our results showed that Kampala could suffer a blow out due to the law of mass action. Therefore, all people in Kampala would suffer an infection rate in which an infection passes within the city and is proportional jointly to the product of the number of people who are infectious, and the number who are at risk of infection.

For the case of mitigation scenarios, washing hands with 50% compliance and regular hand washing of 6 times a day, was the most effective and sustainable to reduce SARS-CoV-2 exposure. This was followed by public wearing of face masks if at least 30% of the population complied, and physical distancing when 20% adhered. Shielding the vulnerable groups was also an effective way of reducing cases of SARS-CoV-2 where relapses were followed by a jump in cases. If schools, bars and churches were opened without compliance to mitigation measures, the highest incidence was observed, leading to a big replacement number. Despite the protective effect of these mitigation measures, evidence shows major challenges in achieving and sustaining compliance to these measures especially the face masks in traditionally non-masking communities (https://apps.who.int/iris/bitstream/handle/10665/329438/9789241516839-eng.pdf?ua=1). Distancing could also be difficult to implement especially in crowded settings such as informal sector workplaces, busy peri-urban settings, in public transportation, among others (http://www.oecd.org/coronavirus/policy-responses/cities-policy-responses-fd1053ff/). On the other hand, achieving hand hygiene through frequent handwashing can be a challenge given the low access to clean water in some settings in Uganda—estimated at less than 50% overall (https://www.unicef.org/uganda/what-we-do/wash). Community mobilization and education to support behavior change is thus necessary to achieve and sustain high levels. This should be accompanied with investments and innovations to expand access to safe water and ensure availability of masks for those who may not afford them (https://www.who.int/emergencies/diseases/novel-coronavirus-2019/question-and-answers-hub/q-a-detail/coronavirus-disease-covid-19-masks).

## Conclusions

This paper demonstrates the benefit of a combination of SARS-Cov2 prevention measures including hand washing, use of masks and social distancing in controlling the pandemic in Uganda. These mitigation measures are most successful when adherence and compliance are practiced. More research is therefore needed to inform the implementation and compliance to these measures across various populations and socioeconomic settings.

### Recommendations

With increasing number of cases, the overarching aim of Uganda should be slowing down the transmission and reducing mortality associated with SARS-Cov2. To achieve this, we recommend that

i. people need to take the risk of infection with SARS-Cov2 seriously and follow the advice issued by national and local health authorities.
ii. although for most people SARS-Cov2 causes only mild illness, it can make some people very ill and fatal especially to older people, and those with preexisting medical conditions (such as high blood pressure, heart problems or diabetes). Protection for this vulnerable population should be mandatory.
iii. sectors and communities should be mobilized to ensure that they take ownership of and participate in the response and in preventing cases through hand washing, public wearing of face masks, and individual-level physical distancing.
iv. sporadic cases and clusters should be controlled to prevent community transmission by continuing to rapid finding and isolating all cases, providing them with appropriate care, and tracing, quarantining, and supporting all contacts.
v. community transmission through infection prevention and control measures should be suppressed, and appropriate and proportionate restrictions on non-essential movement restricted. For high risk cities, a geographical lockdown maybe inevitable.
vi. mortality should be reduced by providing appropriate clinical care for those affected by SARS-Cov2, ensuring the continuity of essential health and social services, and protecting frontline workers and vulnerable populations.

## Data Availability

The data used in the study is readily available on the Ministry of Health website at https://www.health.go.ug, and https://www.who.int.

